# Investigating the Role of Neuroinflammation and Brain Clearance in Frontotemporal Lobar Degeneration using 7T MRI and Biofluid Markers: Protocol for an observational cross-sectional cohort study

**DOI:** 10.1101/2025.03.24.25324503

**Authors:** Fieke A. M. Prinse, Louise van der Weerd, John C. van Swieten, Itamar Ronen, Harro Seelaar, Lydiane Hirschler, Chloé Najac, Elise G. P. Dopper

## Abstract

**Introduction:** Frontotemporal lobar degeneration (FTLD) is the second most common early-onset dementia. Several studies demonstrated that neuroinflammation and iron accumulation occurs in FTLD. However, the timing and relevance of these processes and whether these two are merely cause or consequence remains unclear. Elucidating the role is crucial to assess the rationale for using anti-inflammatory therapies in FTLD. Additionally, the process of glymphatic brain clearance has gained attention as a potential contributor in the disease pathophysiology.

**Methods and analysis:** In this multimodal biomarker study, we use a combination of ultra-high field (7T) MRI, blood, and cerebrospinal fluid (CSF) biomarkers to investigate the role of neuroinflammation, iron accumulation, and brain clearance in FTLD, and to identify biomarkers to differentiate FTLD-TDP from FTLD-tau. We aim to include 25 patients with probable FTLD-tau, 25 with probable FTLD-TDP, and 50 healthy individuals with 50% risk to develop FTLD. We will use several MRI techniques, including magnetic resonance spectroscopy, diffusion weighted spectroscopy, and quantitative susceptibility mapping. In addition, we will assess the prevalence of perivascular spaces and the mobility of CSF to address glymphatic brain clearance. We will compare quantitative MR markers between patients with FTLD-tau and FTLD-TDP, presymptomatic mutation carriers, and healthy controls and correlate these measures with clinical data and biomarkers in blood and CSF.

**Ethics and dissemination:** We obtained ethical approval from the Medical Ethics Committee Leiden Den Haag Delft (NL78272.058.21). The results will be disseminated through presentations at national and international conferences, open access peer-reviewed publications, ClinicalTrials.gov and to the public through social media posts and annual newsletters.

**Trial registration number:** This study is registered on ClinicalTrials.gov (NCT06870838).

**Strengths and Limitations of this study:**

- The first study using 7T MR in the full spectrum of both presymptomatic and symptomatic genetic and sporadic FTLD.
- This study will provide valuable insights in the role of neuroinflammation, iron accumulation, and brain clearance and their role in the disease process of FTLD.
- Multimodal approach using MR and biofluid biomarkers, clinical, neurological, neuropsychological, and neuropsychiatric evaluation.
- No evaluation of the progression of MR and CSF biomarkers due to the cross-sectional design, however, we do perform clinical follow-up.
- The rarity of some specific mutations could result in too small subgroups for sub analyses for each type of mutation separately, but we hope to undermine this issue with the multicenter data inclusion and acquisition.

## 1. Introduction

Frontotemporal lobar degeneration (FTLD) is a neurodegenerative disorder characterized by neuronal loss, gliosis, and pathological protein deposition, most prominently in the frontal and temporal lobes ^1^ ^2^. These pathological changes lead to variable manifestations of cognitive, behavioral, language and motor symptoms (fig. 1) ^3^. Two major subtypes of FTLD have been identified: FTLD with tau accumulation (FTLD-tau) and FTLD with TAR DNA-binding protein 43 aggregates (FTLD-TDP)^2^. About 20-30% of FTLD cases are familial with an autosomal dominant pattern of inheritance. The most commons genetic causes are mutations in *microtubule-associated protein tau* (*MAPT*) ^4^ which are associated with FTLD-tau and mutations in *progranulin* (*GRN*) ^5^ ^6^ or the *chromosome 9 open reading frame 72* hexanucleotide repeat expansion (*C9orf72* HRE) ^7^ ^8^, both associated with FTLD-TDP. The majority of FTLD patients are sporadic, meaning that a definitive pathological diagnosis can only be made with a postmortem neuropathological examination ^9^. However, there are some strong clinicopathological correlations: (sporadic cases of) progressive supranuclear palsy (PSP), corticobasal syndrome (CBS) and non-fluent variant primary progressive aphasia (nfvPPA) are most commonly associated with FTLD-tau, whereas frontotemporal dementia with motor neuron disease (FTD-MND) and semantic variant primary progressive aphasia (svPPA) are typically associated with FTLD-TDP ^10^. Behavioral variant frontotemporal dementia (bvFTD) is the most common clinical presentation of FTLD and associated with both FTLD-tau and FTLD-TDP. Therefore, a reliable distinction between these proteinopathies cannot be obtained during life in sporadic bvFTD cases ^9^. Treatment options are currently under investigation in genetic FTLD, however, no therapies are yet available for this devastating disorder. Considering that potential treatment will most likely be protein-specific ^11^, diagnostic tools to predict the underlying pathology (FTLD-tau from FTLD-TDP) during life are crucial.

**Figure 1:**
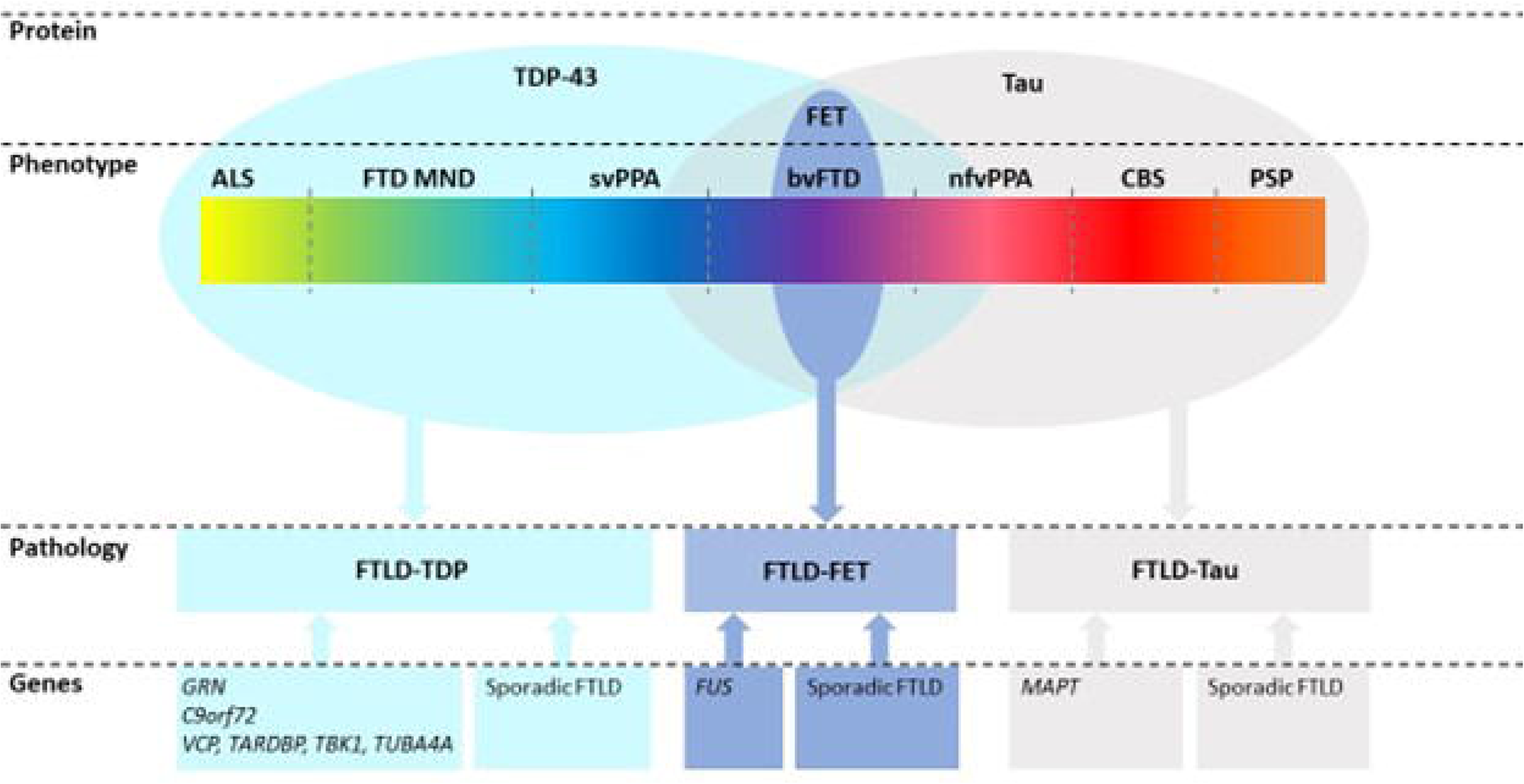
Clinical, pathological, and genetic spectrum of FTLD. Genetic forms have predictable pathology: *GRN* mutations and *C9orf72* HRE show TDP-43 pathology, whereas *MAPT* mutations show tau pathology. In contrast, across the clinical spectrum of FTLD variable underlying pathologies and genetic forms can be found. Abbreviations: ALS, amyotrophic lateral sclerosis; bvFTD, behavioral variant FTD; C9orf72, chromosome 9 open reading frame 72; CBS, corticobasal syndrome; FET, FUS, Ewing sarcoma, and TATA-binding protein-associated factor 15; FTD, frontotemporal dementia; FUS, fused in sarcoma; GRN, progranulin; MAPT, microtubule associated protein tau; MND, motor neuron disease; nfvPPA, non-fluent variant primary progressive aphasia; PSP, progressive supranuclear palsy; svPPA, semantic variant primary progressive aphasia; TARDBP, transactive response DNA-binding protein, TBK-1, TANK-binding kinase 1; tau, tubulin associated unit; TDP-43 transactive response DNA-binding protein 43, TUBA4A tubulin alpha 4A; VCP, vasolin-containing protein. Adapted from Meeter (2017)^3^.

Neuroinflammation, iron accumulation, and brain clearance have (recently) gained a lot of interest. Thus, unraveling their role in the pathophysiological cascade leading to protein accumulation and eventually neuronal loss is crucial for the development of therapeutic targets. In this study, we aim to elucidate the role and timing of these processes in FTLD using a combination of clinical measures, ultra-high field (7T) MRI, and fluid biomarkers.

### 1.1. Neuroinflammation

Neuroinflammation has been recognized as a strong component in both FTLD-tau and FTLD-TDP, as evidenced by the activation of microglia, astrocytes, and increase in proinflammatory cytokines ^12^. However, elucidating the timing of neuroinflammation in the disease process is crucial in order to verify the rationale for applying anti-inflammatory therapies in FTLD patients. We will employ a multimodal MR-based approach, and use a combination of blood and CSF markers, proton Magnetic Resonance Spectroscopy (^1^H MRS), diffusion-weighted ^1^H MRS (^1^H dMRS), and quantitative susceptibility mapping (QSM).

^1^H MRS provides information about cell-specific processes via reporting of the concentration of intracellular metabolites: total N-acetyl-aspartate (tNAA=NAA+NAAG) and glutamate (Glu) primarily, found in (mature) neurons, and myo-inositol (mI) and choline compounds (tCho=Cho+PCho+GPC) in glia cells [Choi 2007]. Increase in mI and tCho indicates glial cell activation or proliferation, a marker for neuroinflammation ^13^ ^14^, and a decrease in tNAA and Glu mirrors loss of intact neurons, i.e., atrophy of neuronal tissue ^14^. Several 1.5T and 3T MRS studies in FTLD have shown increased mI/creatine ratio, and decreased Glu/glutamine and tNAA/creatine ratio in the anterior cingulate cortex and/or the temporoparietal cortex ^14–16^, indicating a decrease in neuronal viability and glial cell activation. Another 3T study in bvFTD, PPA, CBS, and PSP have found comparable results in the prefrontal cortex (i.e., decreased tNAA and Glu levels), which correlated with behavioral impairment ^17^. A single 7T MRS study in FTLD has shown reduced GABA and Glu concentrations, reflecting loss of inhibitory and excitatory neurotransmitters associated with memory and synaptic plasticity ^18^.

^1^H dMRS measures the apparent diffusion coefficient (ADC) of intracellular metabolites and thus provides information about the intracellular microstructural properties of specific cell types ^19–21^. A decrease in the ADC of tNAA might be attributed to neuronal degeneration as shown in Alzheimer’s disease (AD) where the lower ADC (tNAA) in AD patients was attributed to the presence of tau protein neurofibrillary tangles ^22^. In addition, it is hypothesized that dMRS is more sensitive to glial reactivity than MRS; specifically, alterations in the diffusion of tCho and mI have been linked to glial hypertrophy driven by the inflammation process in humans and animal models ^23–25^. Although dMRS has not been used to investigate neuroinflammation in FTLD yet, it has been used in other diseases such as multiple sclerosis (MS)^26^, systemic lupus erythematosus ^13^, and amyotrophic lateral sclerosis (ALS) ^27^.These studies all showed an increase in diffusivity of mI and tCho in the participants with inflammation, pointing to glial activation.

### 1.2. Iron accumulation

Pathological iron accumulation is strongly correlated with neuroinflammation in a number of neurodegenerative diseases, e.g., AD ^28^ ^29^, and Huntington’s disease ^30^. Recent correlative T2*-weighted MRI and histopathological studies in FTLD-tau and FTLD-TDP brain tissue have shown an association between neuroinflammation and brain iron deposition ^31^ ^32^. In samples of FTLD-TDP, iron rich pathology was found in the upper cortical layers together with diffuse cortical speckling, which was not found in FTLD-tau. These cases presented with iron rich pathology in the middle and deep cortical layers and superficial white matter. Additionally, these findings were distinct from AD and healthy controls ^31^. Another study showed cortical iron accumulation associated with the severity of proteinopathies on histology, which was reflected in the T2*-w MRI as well. However, the different patterns of iron accumulation between FTLD-TDP and FTLD-tau were not shown, but this discrepancy might be explained by the different pathological subtypes included in the two studies ^32^. Additionally, three studies investigated iron accumulation in postmortem brains of PSP ^33^, ALS and FTLD at 7T ^34^, and a combination of ALS, FTLD, AD, PSP, vascular dementia and Lewy body dementia also at 7T ^35^ using ultra high field MRI. These studies found an increase of iron load in multiple subcortical, e.g., the claustrum, caudate nucleus, globus pallidus, thalamus, and subthalamus. However, these studies were semi-quantitative and the separation between paramagnetic and diamagnetic substances is difficult. Therefore, in vivo neuroimaging sensitized to iron is a suitable candidate for establishing outcome measures for protein-specific disease progression, reflecting neuroinflammation in FTLD.

In contrast to AD, only one in vivo iron-MRI study has been conducted in FTLD. This study showed an increase in iron content of the bilateral superior frontal and temporal gyri, anterior cingulate, insula, and red nucleus in bvFTD patients compared to controls. Patients with PPA had an increase in iron content in the left superior temporal gyrus compared to healthy controls. Additionally, there was a strong positive correlation between apathy and disinhibition and iron content in the superior frontal gyrus and putamen, respectively ^36^. However, this study had a relatively small sample size and used susceptibility-weighted scans at 1.5 T MRI, resulting in relatively low signal to noise ratio and resolution and an inability to quantify diamagnetic and paramagnetic contributions.

Quantitative susceptibility mapping (QSM) enables quantification of regional brain iron deposition ^37^, based on the correlation between iron concentration in tissue and local phase changes of the MRI signal. An additional benefit from ultrahigh field, the disturbances caused by the presence of iron are detected even better due to increased sensitivity for tissue related magnetic field disturbances ^38^. High resolution postmortem MRI studies demonstrated that QSM is a technique with high sensitivity and specificity for tissue iron content changes in grey matter ^28^ ^39–41^. In the last few years , in vivo QSM has been applied in multiple other neurodegenerative or neuroinflammatory diseases, such as Parkinson’s Disease ^42^, Parkinsonisms ^43^, ALS ^44^, Systemic Lupus Erythematosus ^45^, and AD ^46^, demonstrating the feasibility of this technique for patient studies; so far FTLD has not been studied in vivo.

### 1.3. Brain clearance

In healthy brains, the flow of CSF within perivascular spaces and its exchange with the interstitial fluid (glymphatic system) has been proposed to help the clearance of waste and toxic proteins ^47^. Aquaporin-4 (AQP4) water channels located on the astrocytic end feet have been shown to facilitate this process ^48^. A potential indicator of dysfunction of the brain clearance is the presence of enlarged PVS, which may result from obstructed waste products ^49–51^. Deficient glymphatic clearance is linked to the accumulation of toxic proteins associated with several neurodegenerative diseases, such as amyloid-beta in AD and cerebral amyloid angiopathy (CAA) ^52^ and alpha-synuclein in PD ^53^.

The role of the glymphatic brain clearance system in FTLD is yet to be determined. A few studies provided some evidence for its implication in FTLD. One study showed elevated levels of AQP4 in the CSF in FTLD patients compared to healthy controls, suggesting reduced glymphatic flow ^54^. Three studies explored the diffusion tensor image analysis along the perivascular space (DTI-ALPS) index in FTLD, which has been proposed as a method to evaluate the glymphatic system ^55–57^. Symptomatic mutation carriers had a lower DTI-ALPS index compared to healthy controls, and this lower index correlated with worse disease severity ^55^ or motor and cognitive functions ^56^. Moreover, a study from the Frontotemporal Lobar Degeneration Neuroimaging Initiative found changes in choroid plexus volume, in blood-oxygen-level-dependent (BOLD) - CSF coupling and in DTI-ALPS index, altogether hinting towards glymphatic system alterations ^57^.

In this study, we will probe the glymphatic system by assessing whether the dynamics of CSF (i.e., the carrier of waste products) change in FTLD patients. To this aim, we will use a high-resolution and CSF-specific MRI-technique called CSF-STREAM (CSF-selective-T_2_-prepared Readout with Acceleration and Mobility Encoding) ^58^, providing detailed whole-brain imaging of CSF-mobility, from subarachnoid spaces down to the level of PVS. Additionally, we will assess the prevalence of enlarged PVS using a high-resolution T_2_-weighted scan.

### 1.4 Study aims

In this multimodal cross-sectional study with one-year clinical follow-up, we will use 7T MRI combined with clinical, neuropsychological, blood, and CSF markers in presymptomatic mutation carriers, symptomatic participants, and healthy controls to investigate neuroinflammation, neurodegeneration, iron accumulation, and brain clearance in FTLD. Furthermore, we aim to identify new biomarkers to differentiate FTLD-tau from FTLD-TDP and to assess the feasibility to predict disease progression. If successful, this will be essential to specifically identify patient-groups and evaluate response to therapy in future clinical trials. The objectives of our study are:

1. To elucidate the role and timing of neuroinflammation and iron accumulation in FTLD by using a combination of clinical measures, 7T MRI, and fluid biomarkers;
2. To differentiate FTLD-TDP and FTLD-tau during life using biomarkers for neuroinflammation;
3. To identify biomarkers to predict and monitor disease progression in FTLD;
4. To explore the role of brain clearance in the disease process of FTLD.

## 2. Methods and analysis

### 2.1 Study design

This is an observational cross-sectional cohort study with a multimodal design and a clinical one-year follow-up. At baseline, the study will involve the following procedures: clinical assessment including neurological and neuropsychological investigation, blood sampling, and a voluntarily lumbar puncture on the first day at the Erasmus MC University Medical Center, (EMC) and two sessions of 7T MRI scanning on the second day at the Leiden University Medical Center (LUMC). After one-year, clinical assessment and blood analyses will be repeated in the EMC to assess disease progression. An overview of the study design is shown in fig. 2. The study started in July 2023 and is expected to be completed in August 2026.

**Fig 2.**
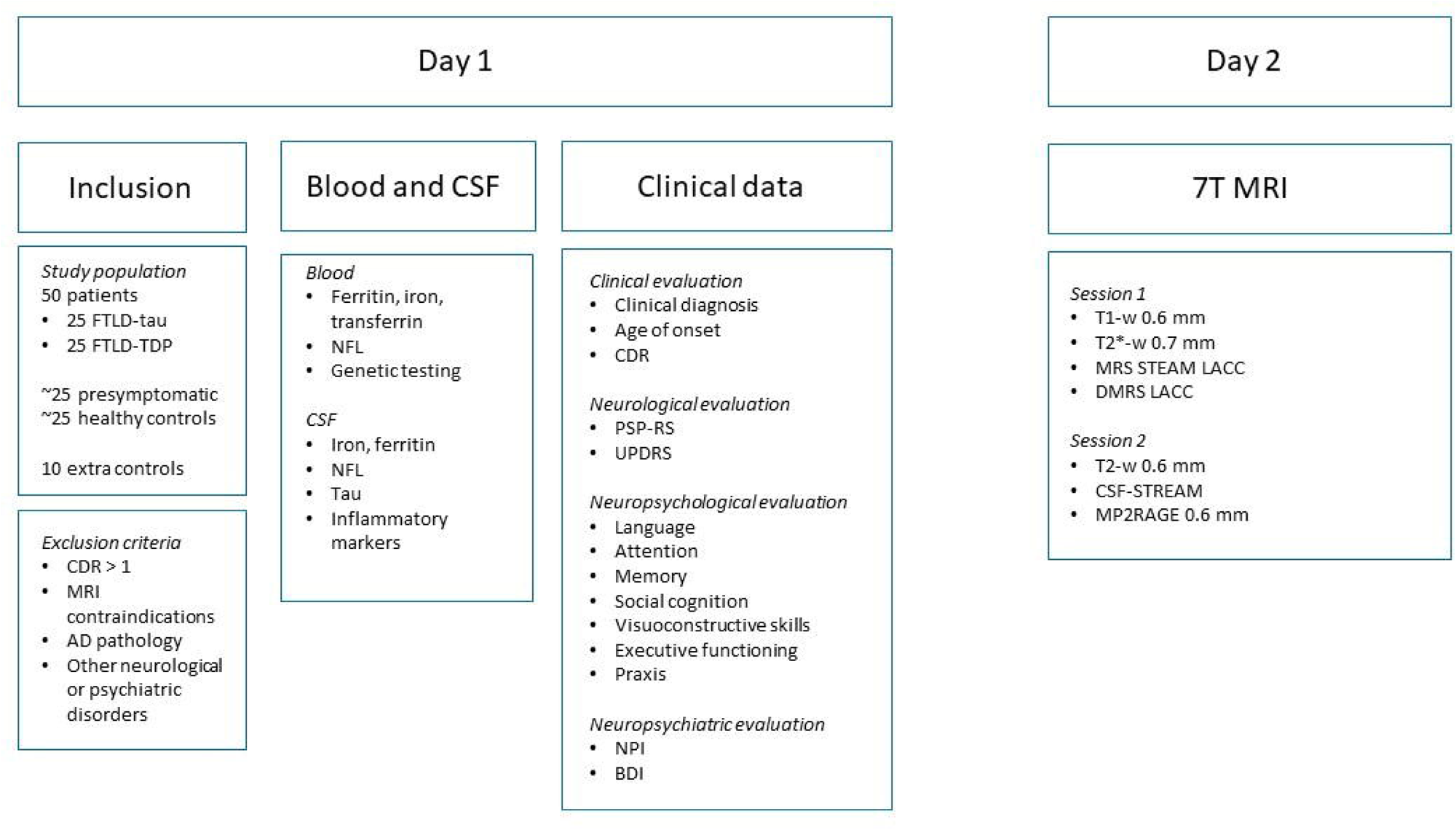
Flowchart of the study design. Clinical data and blood collection will be repeated after 1 year of follow-up. Abbreviations: CSF-STREAM: CSF-selective-T2 prepared Readout with Acceleration and Mobility Encoding. CDR: clinical dementia rating. DMRS: Diffusion weighted spectroscopy. FAB: Frontal assessment Battery. FTLD: frontotemporal lobar degeneration. LACC: Lateral anterior cingulate cortex. MOCA: Montreal Cognitive Assessment. MRS: Magnetic resonance spectroscopy. NFL: neurofilament light chain. NPI: Neuropsychiatric inventory. PSP-RS: Progressive supranuclear palsy rating scale. UPDRS: Unified Parkinson’s Disease Rating Scale.

### 2.2. Ethics and dissemination

This study is registered on ClinicalTrials.gov (NCT06870838). The study was approved by the Medical Ethics Committee Leiden Den Haag Delft (NL78272.058.21). All participants are asked for written informed consent before inclusion in accordance with the World Medical Association Declaration of Helsinki.

The study findings will be presented at (inter)national conferences and shared through publication in international open access peer-reviewed journals and on ClinicalTrials.gov.

### 2.3. Population

Participants will be included at the neurology department of the EMC. We aim to include 25 patients with probable or definite FTLD-tau, 25 patients with probable or definite FTLD-TDP, 50 healthy individuals with 50% risk to carry a mutation in *MAPT* or *GRN,* or the *C9orf72* HRE. If necessary for age matching, we will include additional 10 healthy subjects without increased risk of FTLD.

Patients with probable FTLD-tau are defined as a clinical diagnosis of PSP, CBS, or nfvPPA, or any clinical FTLD spectrum diagnosis with a proven *MAPT* mutation. In patients with CBS or nfvPPA we will use CSF analyses and genetic screening to rule out underlying AD pathology or having a pathogenetic variant causing FTLD-TDP pathology. Patients with probable FTLD-TDP are indicated by either a clinical diagnosis of svPPA, FTLD-ALS or any clinical FTLD spectrum diagnosis with a proven *GRN* mutation or *C9orf72* HRE. Clinical diagnoses are made according to current clinical criteria ^59–62^.

At-risk individuals will be recruited through our ongoing FTD-RisC project ^63^. These individuals have no clinical signs of an FTLD phenotype but have a first degree relative with a *MAPT or GRN* mutation or *C9orf72* HRE. These subjects therefore all have a 50% risk to carry one of these genetic variants and develop FTLD later in life. Through anonymous genetic screening, this group will be divided into presymptomatic mutation carriers and healthy individuals without a mutation (i.e., healthy controls). Since these at-risk individuals are expected to be younger than the patient groups, 10 additional healthy individuals, for instance spouses of patients, might be included for age-matching.

Exclusion criteria include the presence of other neurological or psychiatric disorders that may affect cognitive function, such as brain tumors, multiple sclerosis, drug or alcohol abuse, or use of psychoactive medications. Other exclusion criteria are CSF markers suggestive of AD, a clinical dementia rating (CDR) score higher than 1, or a contra-indication to undergo MRI.

### 2.4. Patient and public involvement

Patients or public were not actively involved in this study design. However, through our ongoing FTD-Risc study since 2010 we are in close contact with FTLD patients and their family members and we have broad experience with similar studies in these patient groups. We inform all participants about study results through an annual newsletter and biannual informative meetings. The broader public will be informed through patient associations and social media.

### 2.5. Clinical data

We will collect basic demographic information including age, sex, and years of education. Furthermore, we will take medical history and neurological examination, including the MDS-Unified Parkinson’s Disease Rating Scale (MDS-UPDRS) in case of signs of parkinsonism. Clinical disease severity will further be evaluated using the CDR scale. Neuropsychological assessment will include Montreal Cognitive Assessment (MOCA), frontal assessment battery (FAB), cognitive screening tools, and tests for language (Boston Naming Task, category and letter fluency), attention, executive functioning (trail making test, Stroop, Brixton, Hayling), memory (15 Words Test, Benson Copy), visuoconstructive functioning (Royall Clock Drawing), social cognition (Emotion Recognition Tasks, Hinting task, Social Interaction Vocabulary Task), and praxis (van Heugten Task for Apraxia and Goldenberg Ideomotor Apraxia Test). Neuropsychiatric symptoms will be assessed with Neuropsychiatric Inventory (NPI) and Beck’s Depression Inventory (BDI).

### 2.6. Blood and CSF

Blood sampling and lumbar punctures will be performed according to clinical standards by experienced physicians. Lumbar punctures are not mandatory in this study unless it is necessary for diagnostics purposes, and thus will only be performed in participants who give additional consent for this. We will collect CSF in polypropylene tubes. Blood and CSF will be centrifuged, and stored in 0.5 mL aliquots at - 80 °C. Blood and CSF samples will be analyzed in the Amsterdam UMC laboratory for the following markers: iron and ferritin for iron accumulation, neurofilament light chain and total tau for neuronal damage, and YKL-40, TREM-1, TREM-2, IL-1, IL-6, TNF-α, GFAP, CHIT1 as inflammatory markers, and AQP4, tau, and TDP-43 in CSF as markers for brain clearance (Table 1).

**Table 1:**
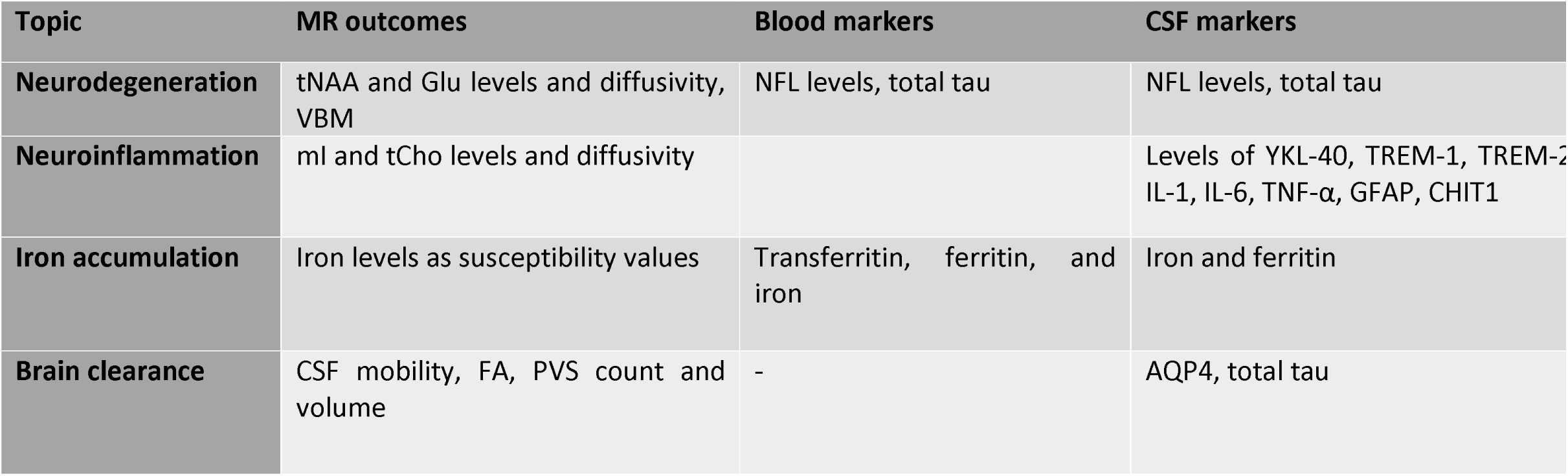
Overview of the acquired biomarkers. Abbreviations: AQP4: aquaporin-4; CHIT1: chitinase 1; CSF: cerebrospinal fluid; FA: fractional anisotropy; GFAP: glial fibrillary acidic protein; Glu: glutamate; IL-1: interleukin-1; IL-6: interleukin-6; mI: myo-inositol; NFL: neurofilament light; tau: tubulin associated unit; tCho: total choline TDP43: transactive response DNA binding protein 43; TNF-α; tumor necrosis factor α; tNAA: total N-acetylaspartate; TREM-1: Triggering receptor expressed on myeloid cells 1; TREM-2: Triggering receptor expressed on myeloid cells 2 ; VBM: voxel-based morphometry; YKL-40: Chitinase-3-like protein 1.

### 2.7. Genetic analysis

In all participants that have not undergone genetic testing in clinical practice, blood samples will be used to screen for genetic causes of FTLD. Participants from families with a known genetic defect will be selectively screened for that type of mutation. In suspected sporadic cases, next generation sequencing including *C9orf72* will be performed to investigate all known genetic causes of FTLD. The genetic testing will only be done at the baseline visit and will be performed anonymously. Genetic screening will be carried out at the clinical genetics’ laboratory of the EMC.

### 2.8. MR acquisition

All scans will be performed at the LUMC using a whole-body human Philips Achieva 7T MR scanner (Philips Healthcare, Best, The Netherlands) with a quadrature transmit and 32-channel receive head coil (Nova Medical, Wilmington, MA, USA). The *first scan session* will consist of a 0.6 mm isotropic 3D MPRAGE T_1_-weighted scan for anatomical information. Then, a whole brain multi-echo gradient echo scan will be acquired with a 0.7 mm isotropic resolution for the QSM analysis. For quantifying metabolites’ level and diffusion, we will use a single-volume ^1^H STEAM sequence (MRS) and a single-volume ^1^H diffusion-weighted semi-LASER sequence (dMRS), respectively. All MRS and dMRS data will be collected in a volume-of-interest (VOI) located in the lateral anterior cingulate cortex. For patients, the VOI will be placed in the hemisphere that is thought to be the most affected according to the symptoms. For healthy controls and at-risk individuals, the VOI will be randomly assigned to the left and the right side. Cardiac triggering is used for dMRS acquisition, using a peripheral pulse unit (Philips, Best, The Netherlands). The *second scan session* will consist of a 0.6 mm isotropic 3D TSE T_2_-weighted scan, a 0.7 mm isotropic MP2RAGE scan, and the 0.45 mm isotropic CSF-STREAM sequence ^58^. As brain clearance is thought to be influenced by respiration and cardiac pulsations, the participants will wear a respiration belt and a peripheral pulse heart monitor unit (Philips Healthcare, Best, The Netherlands) during those scans ^48^ ^64^. Detailed information for the scan parameters is given in Table 2.

**Table 2:**
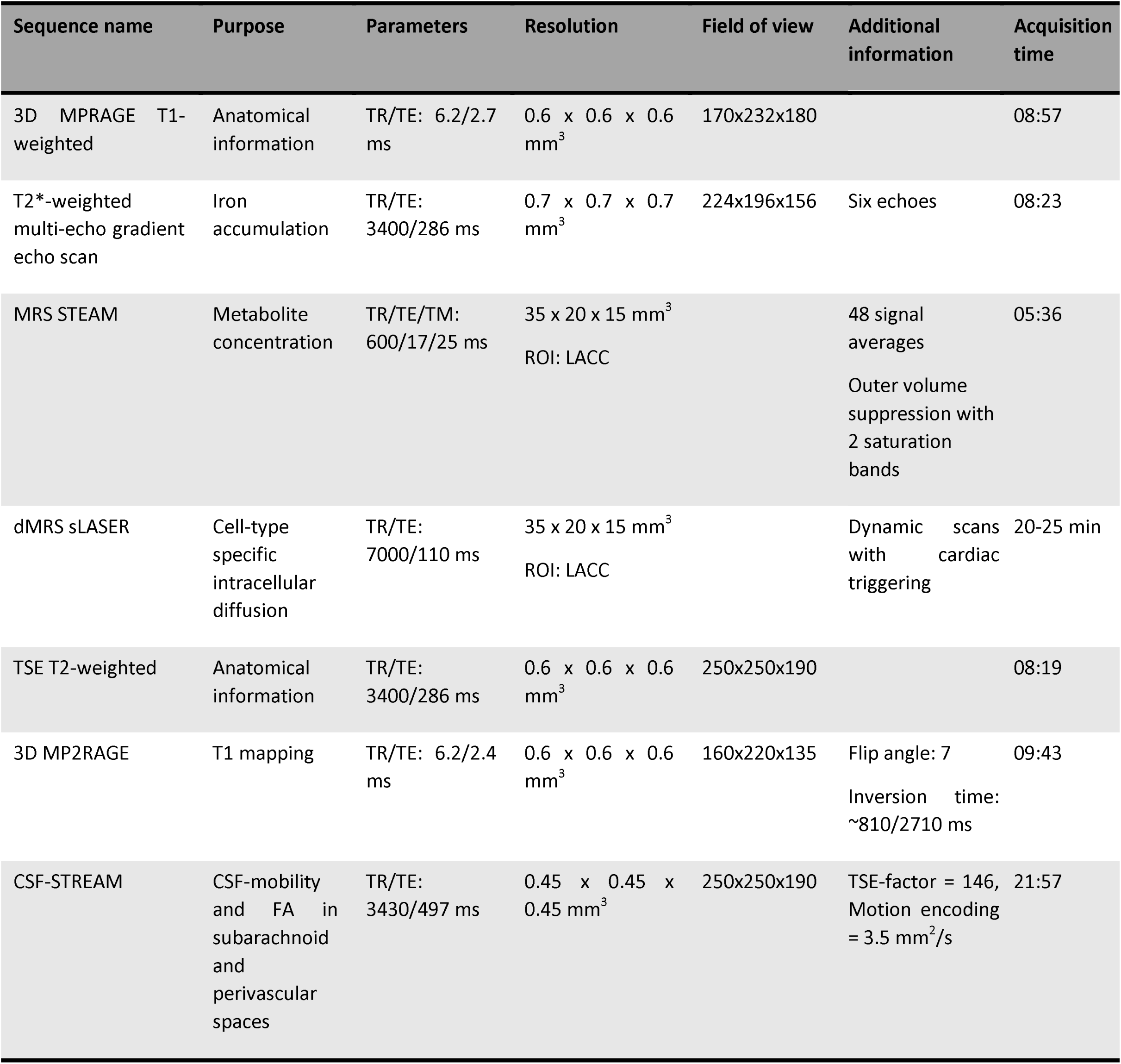
Standardized scan protocol on the 7T MRI scanner. Abbreviations: LACC, lateral anterior cingulate cortex.

### 2.9. MR data processing

Established pipelines for basic segmentation of anatomical images, co-registration, and basic motion and intensity corrections will be applied using FSL ^65^ and SPM ^66^. Voxel-based morphometry will be used to quantify brain volume. We will use the QSM processing pipeline toolbox SEPIA ^67^ to generate QSM maps from multi-echo data. From the maps, we will obtain voxel-wise susceptibility values indicative of iron content in predefined regions of interest in affected brain areas, e.g., the frontal, temporal, and parietal cortex, and the basal ganglia. The exact regions will depend on the quality and signal-to-noise ratio of the obtained data. MRS and dMRS data processing will be performed with in-house MATLAB pipelines. Levels of brain metabolites, e.g., tNAA, mI, tCho, Glu and tCr, will be quantified using LCModel^68^. ADCs for the various metabolites, measured with dMRS, will be calculated as shown previously ^20^. The CSF-STREAM scans will be analyzed using in-house MATLAB and python pipelines. We will extract CSF-mobility and fractional anisotropy (FA) values in PVS around penetrating vessels as well as in subarachnoid spaces around larger vessels. PVS burden will be determined on high-resolution T2-weighted scans.

### 2.10. Power calculations

The study population is based on the sample size calculations for the main MR outcome measures. To test the sample size needed to identify differences in these outcome measures, the software G*Power was used ^69^. The aimed power was set at 0.8 and the significance level α at 0.05. For QSM the power calculation was based on changes in iron content observed in Huntington’s Disease using QSM ^70^. Assuming two group, the caudate nucleus of healthy controls is estimated to have a susceptibility value of 0.043 ± 0.019 parts per million (ppm). Patients with Huntington’s Disease are estimated to have a susceptibility value of 0.059 ± 0.016 ppm. This results in an effect size of 0.9 and a required sample size of at least 21 subjects per group. The power calculation for MRS was based on data from a similar 7T project in AD ^71^. For tCho/tCr concentration the mean was 0.19 ± 0.02 in healthy controls and 0.22 ± 0.03 in AD and for Ins/tCr, the mean was 0.74 ± 0.06 in controls and 0.87 ± 0.10 in AD, resulting in a required sample size of at least 10 to 16 subjects per group for detecting a 10% group difference. A full-fledged power calculation for dMRS experiments can be found in ^13^. In short, based on experimental conditions similar to our study, 11-13 subjects are required in each group to detect a group difference of 10% in the diffusion of the intraneuronal marker NAA.

### 2.11. Statistical analyses

All main outcome measures will be tested for normal distributions, and if applicable, non-parametric tests will be used. For all statistics, the level of significance is set at p < 0.05, with Bonferroni correcting for multiple testing. Clinical data will be presented as continuous or categorical values, depending on the questionnaire or cognitive tests. Markers for neurodegeneration and neuroinflammation in blood and CSF and MR markers will be expressed as continuous variables. MR markers will include quantitative susceptibility values from the QSM, metabolites’ absolute concentrations and ratios to total creatine for MRS, metabolites’ ADCs for dMRS, and CSF-mobility and FA for the CSF-STREAM data.

Demographics (sex, age, level of education), clinical data (i.e., CDR, MDS-UPDRS, neuropsychological test results), and blood and CSF markers will be compared between groups using one-way ANOVA’s. We will use regression analyses to investigate group differences (symptomatic participants vs presymptomatic mutation carrier’s vs controls and FTLD-TDP vs FTLD-tau) in the various MR markers. Sex, age, and brain tissue volume will be used as covariates in these analyses. Furthermore, we will correlate the various MR markers with each other and with clinical, blood and CSF measures.

## 3. Discussion

FTLD is a neurodegenerative disorder that leads to various clinical manifestations, all with major impact on the life of patients and their relatives. There is currently no disease-modifying therapy available. In light of the development of treatment strategies, deeper knowledge of the exact mechanism is crucial. Additionally, distinction between FTLD-tau and FTLD-TDP during life would be very valuable for enrollment in therapeutic trials, since it is assumed that the disorder requires protein-specific targeted treatment strategies. The involvement of neuroinflammation in FTLD is an established hypothesis and would be a valuable testing point for drug development. However, the exact mechanism and timing relative to neurodegeneration remain ambiguous and can depend on the specific genetic and pathological factors involved. Therefore, in this study, we will investigate the role and combination of neuroinflammation, neurodegeneration, iron accumulation, and brain clearance in FTLD.

7T MR has promising potentials in visualization of neuroinflammatory processes and development of biomarkers in FTLD ^38^ ^72^ ^73^. Firstly, 7T MR provides an improved spatial and spectral resolution compared to lower field strengths, allowing more precise visualization of cell-specific metabolites, small brain structures and regions affected by neuroinflammation. Secondly, the increased signal-to-noise-ratio enables better detection of subtle changes in brain tissue, allowing early identification of neuroinflammation. Thirdly, 7T MRI allows for more detailed imaging of cerebral microvasculature. Neuroinflammation often involves alterations in the blood-brain-barrier and microvascular changes, which can be better characterized at this field strength. The combination of these advantages allows to map iron accumulation with quantitative susceptibility mapping, visualize microstructural changes in metabolites using d(MRS), and explore brain clearance through PVS volume quantification and CSF-mobility assessment in FTLD. These imaging findings will be correlated for validation with established fluid biomarkers and neurological and neuropsychological assessments.

In addition to the abovementioned sequences, we acquire an additional high-resolution MP2RAGE scan for T1 mapping ^74^ ^75^. One study showed altered microstructural changes in the striatum of PD patients presented as degeneration of parts of the putamen using quantitative MRI mapping of T1w/T2w ratios^76^. As T1-weighted scans and T2-weighted scans are easily available to the clinic, this could help to evaluate the link between brain microstructural changes and symptoms in neurodegenerative disease, without the need of complicated scans ^76^.

Our study design exhibits several strengths. Firstly, abundant MRI studies have already been performed in FTLD ^77^, but studies using 7T MRI are sparse. 7T MRI offers a gain in sensitivity and specificity, which will significantly boost spatial and spectral resolution on our MRI and (d)MRS scans. Secondly, we will include sporadic and genetic participants with both (suspected) FTLD-tau and FTLD-TDP pathology. This will allow us to cover a wider spectrum of the disease within the same study, as opposed to the current published studies that have either focused on genetic FTLD or only specific clinical subtypes. Particularly, we will be able to compare FTLD-tau and FTLD-TDP and potentially identify markers or test to use for ante mortem definite diagnosis. Thirdly, we will include at risk individuals and symptomatic participants, allowing us to study the timing of the disease’s mechanisms. Lastly, for this study we will collaborate with the FTD center of the University of Pennsylvania, Philadelphia. They perform a similar study, with different specific research questions. We have harmonized MRI acquisition protocols and signed a data transfer agreement, thus data from both sites can be used for both purposes. The two sites make use of MRI scanners from different vendors. While protocols are harmonized as much as possible, they might vary slightly, immediately providing us with the opportunity to test the robustness of our potential biomarkers.

One limitation of our study is that FTLD is a heterogeneous disease with various and unclear mechanisms leading to the disease. There is a possibility that subgroups become smaller than expected. We will try to undermine this problem by using the multicenter approach as both sites will include independently. Therefore, mutations or pathology that might be rarer at one site can be resolved by the inclusion at the other site. Another limitation is the cross-sectional design of the study. Longitudinal data would be ideal to evaluate the abovementioned biomarkers for progression of the disease. This is not possible in our current design, however, this could be evaluated in the future.

In conclusion, our study aims at disentangling the role of neurodegeneration, neuroinflammation, iron accumulation and brain clearance throughout the course of the disease in FTLD. For this purpose, we use a unique combination of clinical data, neuropsychological investigation, 7T MRI, and fluid biomarkers in both the presymptomatic and symptomatic disease stage and in both sporadic and genetic FTLD. To our knowledge, this is the first study to integrate these approaches using various MR sequences at 7T.

## Data Availability

No data have been used in this manuscript.

## Ethics statements

### Patient consent for publication

Not Applicable

### Data sharing statement

No data was involved in the writing of this paper.

### Author Contributions

ED conceptualized and designed the protocol including methodology with the help of IR, LvdW, JvS, LH, and CN. Initial funding acquisition was obtained by ED and LH. ED and FP are the current study coordinators, responsible for project administration, organizing resources, participant inclusion, and data curation. Supervision is done by ED, CN, LH, and LvdW. Technical imaging aspects are supervised by CN, LvdW, IR, and LH. FP made the original draft of this paper. LvdW, JvS, IR, HS, LH, CN, and ED reviewed and approved all drafts.

## Acknowledgement and sources of funding

This study is supported by Alzheimer Nederland (WE.06-2021-01) (ED) and the Joint Program for Neurodegenerative Diseases (JPND) on (MINDFACE - 113055) (JvS). LH is funded by the European Union’s Horizon 2020 Research and Innovation Programme under grant agreement No. 825664 and by a grant from the Leducq Foundation and the Leducq Foundation for Cardiovascular Research (23CVD03).

Two of the authors of this publication are members of the European Reference Network for Rare Neurological Diseases - Project ID No 739510.

